# The Role of Mitochondrial genome abundance in Alzheimer’s Disease

**DOI:** 10.1101/2022.05.27.22275698

**Authors:** Nadia V. Harerimana, Devashi Paliwali, Carmen Romero-Molina, David A. Bennett, Judy Pa, Alison Goate, Russell H. Swerdlow, Shea J. Andrews

## Abstract

Mitochondrial dysfunction is an early and prominent feature of Alzheimer’s disease (AD), with impaired energy metabolism preceding the onset of clinical symptoms. Here we propose an update to the mitochondrial dysfunction hypothesis of AD based on recent results examining the role of mitochondrial genome abundance in AD. In a large post-mortem study, we show that lower brain mitochondrial genome abundance is associated with a greater odds of AD neuropathological change and worse cognitive performance. We hypothesize that lower mitochondrial genome abundance impairs mitochondrial function by reducing mitochondrial bioenergetics, thereby impacting neuronal and glial cell function. However, it remains to be determined if mitochondrial dysfunction causes, mediates, or is a by-product of AD pathogenesis. Additional support for this hypothesis will be generated by linking peripheral blood mitochondrial genome abundance to AD and establishing clinical trials of compounds that upregulate total mitochondrial genome abundance or boost mitochondrial mass.

**RESEARCH IN CONTEXT:** *Systematic Review:* The authors used PubMed to review the literature on mitochondrial genomics in Alzheimer’s disease (AD) using the following search term: *mitochondria* AND (“copy number” OR heteroplasmy OR haplogroup*) AND* “*Alzheimer’s disease*”. The accumulated evidence suggested that increased mitochondrial genome abundance is neuroprotective, but found conflicting evidence for the association of mitochondrial heteroplasmy or specific haplogroups with AD.

*Interpretation:* We found that higher mtDNA abundance was robustly associated with reduced AD neuropathology burden and higher neurocognitive performance. Given these findings, we propose an updated hypothesis for mitochondrial dysfunction in AD: that mitochondrial genome abundance is a relevant mechanism in AD pathogenesis. We postulate that baseline mtDNA abundance itself contributes to baseline mitochondrial function and lifetime risk, and that propensity and sensitivity to mtDNA depletion further modulate risk, histopathology, and clinical decline.

*Future directions:* Using statistical genetics approaches, examining the association of peripheral mtDNA abundance with AD, and upregulating mtDNA abundance, would further strengthen the evidence of a causal role for mtDNA abundance and mitochondrial dysfunction in AD pathogenesis

## 1. NARRATIVE

### 1.1 Contextual background

Alzheimer’s disease (AD), a debilitating neurological condition characterized by memory deficits and cognitive and behavioral impairment, affects more than 43.8 million people worldwide [1]. Attempts to identify therapeutics preventing or delaying the onset of AD have focused primarily on the role of amyloid-β peptide and hyperphosphorylated tau, which are the classical neuropathological hallmarks of AD [2]. The limited success to date in identifying disease-modifying therapies has led to research exploring other potential mechanisms underlying disease pathogenesis such as mitochondrial dysfunction.

Mitochondria are intracellular organelles involved in fundamental cellular processes, including oxidative phosphorylation, apoptotic signaling, regulation of innate and adaptive immunity, and calcium storage [3]. Mitochondria have their own circular genome (mtDNA), which is maternally inherited and encodes proteins crucial for the electron transport chain. Due to the integral role of mitochondria in cellular processes, mitochondrial dysfunction has been reported in several age-related [4] and neurodegenerative diseases [5]. Mitochondrial dysfunction is an early and prominent feature of AD with impaired energy metabolism preceding the onset of clinical symptoms and mitochondria in AD patients exhibiting altered mitochondrial dynamics, disrupted bioenergetics, and increased oxidative stress [5]. Consequently, these early findings have contributed to the formulation of the mitochondrial cascade hypothesis, which proposes that one’s genetically determined baseline mitochondrial function subsequently declines at a genetically determined rate as the brain ages under the influence of its environmental background. The hypothesis further proposes that the traditional AD chronology is initiated upon surpassing a threshold of age-associated mitochondrial decline [6].

As a multicopy genome, mtDNAs are present as many copies per cell, with mtDNA copy number - mtDNA abundance - serving as a biomarker of mitochondrial activity that can be used as a proxy for mitochondria function [7,8]. The development of new methods for robustly estimating mtDNA abundance [9] and calling mtDNA genetic variation has allowed us to evaluate mitochondrial dysfunction at the population level and investigate the link between mitochondrial dysfunction and AD pathogenesis. Here, we evaluate association of mtDNA abundance in relation to AD neuropathology and cognitive function and we propose an updated hypothesis for mitochondrial dysfunction in AD - that mtDNA abundance is a relevant mechanism in AD pathogenesis

### 1.2 Mitochondrial dysfunction in AD

Neuritic plaques and neurofibrillary tangles are the core neuropathological hallmarks of AD. However, no therapies targeting the ‘amyloid cascade’ have been successful in preventing or ameliorating the AD pathogens [2]. The lack of success of Aβ targeting therapies and inconsistent evidence of the causal role of Aβ in sporadic AD has prompted criticism of the amyloid cascade model [10]. The heterogeneity in AD clinical symptoms and onset indicates that other factors influence disease pathogenesis. AD research is now expanding to investigate the role of multiple pathways that could explain or contribute to AD [10]. Mitochondrial dysfunction is one such avenue with a growing body of robust evidence implicating its role in neurodegeneration.

Several mitochondrial processes are dysregulated or dysfunctional in AD, such as disrupted bioenergetics, increased oxidative stress, and altered genomic homeostasis. Glucose hypometabolism, measured using fluorodeoxyglucose-positron emission tomography is a surrogate marker of energy metabolism in the brain, with lower glucose uptake generally interpreted as impaired energy metabolism through oxidative phosphorylation. Glucose hypometabolism is a consistent feature of AD and is observed in the parietotemporal association area, posterior cingulate, and precuneus 10-15 years before the clinical onset of symptoms [11]. While glucose metabolism is influenced by mitochondrial function, it may not accurately quantify mitochondrial function. 18F-BCPP-EF is a recently developed radioligand for mitochondrial complex I abundance and is reduced in the parahippocampal cortex of early-stage living AD patients [12] and coincides with tau deposition in the transentorhinal and entorhinal regions [13].

Mitochondria in AD brains exhibit morphological changes resulting from dysregulated fusion and fission, such as lower numbers, abnormal sizes, and shapes; and the internal membrane has reduced and broken cristae. Biomarkers of oxidative stress, such as reactive oxygen species, are increased in the brains of individuals with AD and are also associated with mtDNA mutations, damage to the mitochondrial respiratory chain, altered mitochondrial membrane permeability, and structure, and disturbed Ca2+ homeostasis [14]. Additionally, multi-omic gene and protein expression studies also support the role of mitochondrial pathways in AD. Modules of co-expressed genes related to mitochondrial function positively correlate with histopathological β-amyloid burden, cognitive decline, and AD clinical diagnosis [15]. Several proteomic profiling studies on AD brains and cerebrospinal biomarkers show modules of co-expressed protein families strongly linked to mitochondrial metabolism [16–18]. RNA sequencing studies of AD brains have found significantly reduced expression of all mtDNA OXPHOS genes, the master mitobiogenesis regulator PGC-1α, and mtRNA stabilizing protein LRPPRC [19].

### 1.3 Study design and main results

The mitochondrial genome contains multiple copies per cell, with the number depending on the bioenergetic needs of each tissue and cell type (Figure 1). Previous studies have shown that mtDNA abundance associates with AD, with lower levels of mtDNAcn from post-mortem brain tissue [20,21] correlating with worse CDR scores [22]. The present study investigated if reduced mtDNAcn within the brain was associated with an increased risk of developing AD using data from post-mortem brain tissue. We found that increased mtDNAcn levels were associated with reduced odds of neuropathological diagnosis of AD and neurofibrillary tangle burden but not with neuritic plaque burden (Table 3, S1). We further examined the association of mtDNAcn levels with additional quantitative measures of AD pathology and cognitive function. Higher mtDNAcn was associated with a lower global AD pathology and tau tangle density and better cognitive performance in domains associated with working memory, episodic memory, semantic memory, perceptual speed, perceptual orientation, and global cognition (Table 4, S4). These results are consistent with the notion that higher mtDNAcn levels reflect healthier mitochondria in the elderly brain.

**Figure 1:**
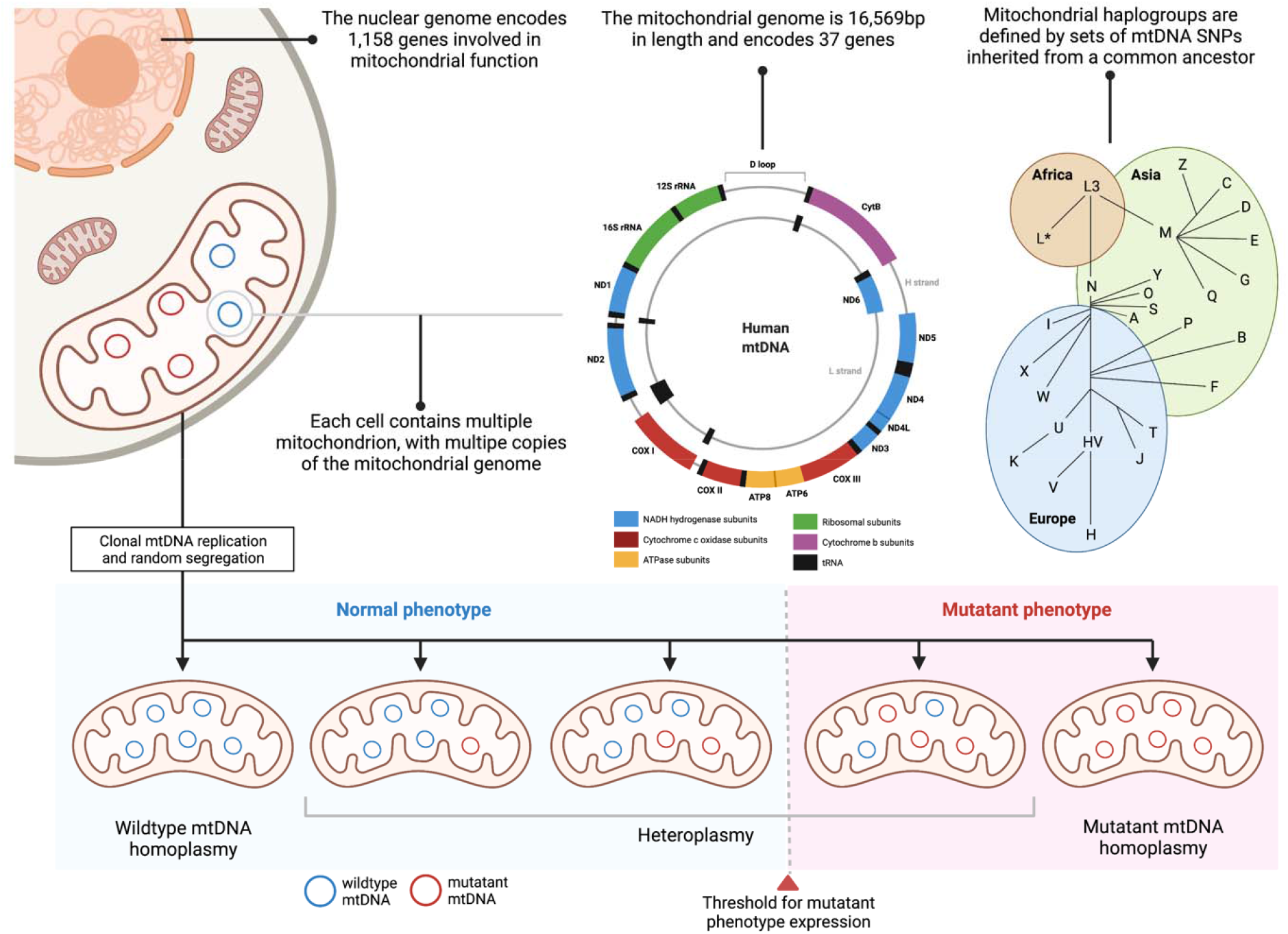
Schematic representation of mitochondrial genomics. Mitochondria are intracellular organelles that contain their own circular genomes. One type of common genetic variation in the mitochondrial genome is characterized as ***mitochondrial haplogroups*** defined by shared sets of mtSNPs that arose during prehistoric human migrations. The mitochondrial genome is a multicopy genome, with ***mitochondrial copy number*** varying depending on the bioenergetic needs of a given cell type. Mutations in the mtDNA can either affect all (***homoplasmy***) or only a fraction of the mtDNAcn molecules (***heteroplasmy***). mtDNA mutations that cause a defect in the OXPHOS system are tolerated until they exceed a certain biochemical threshold.

**Figure 2:**
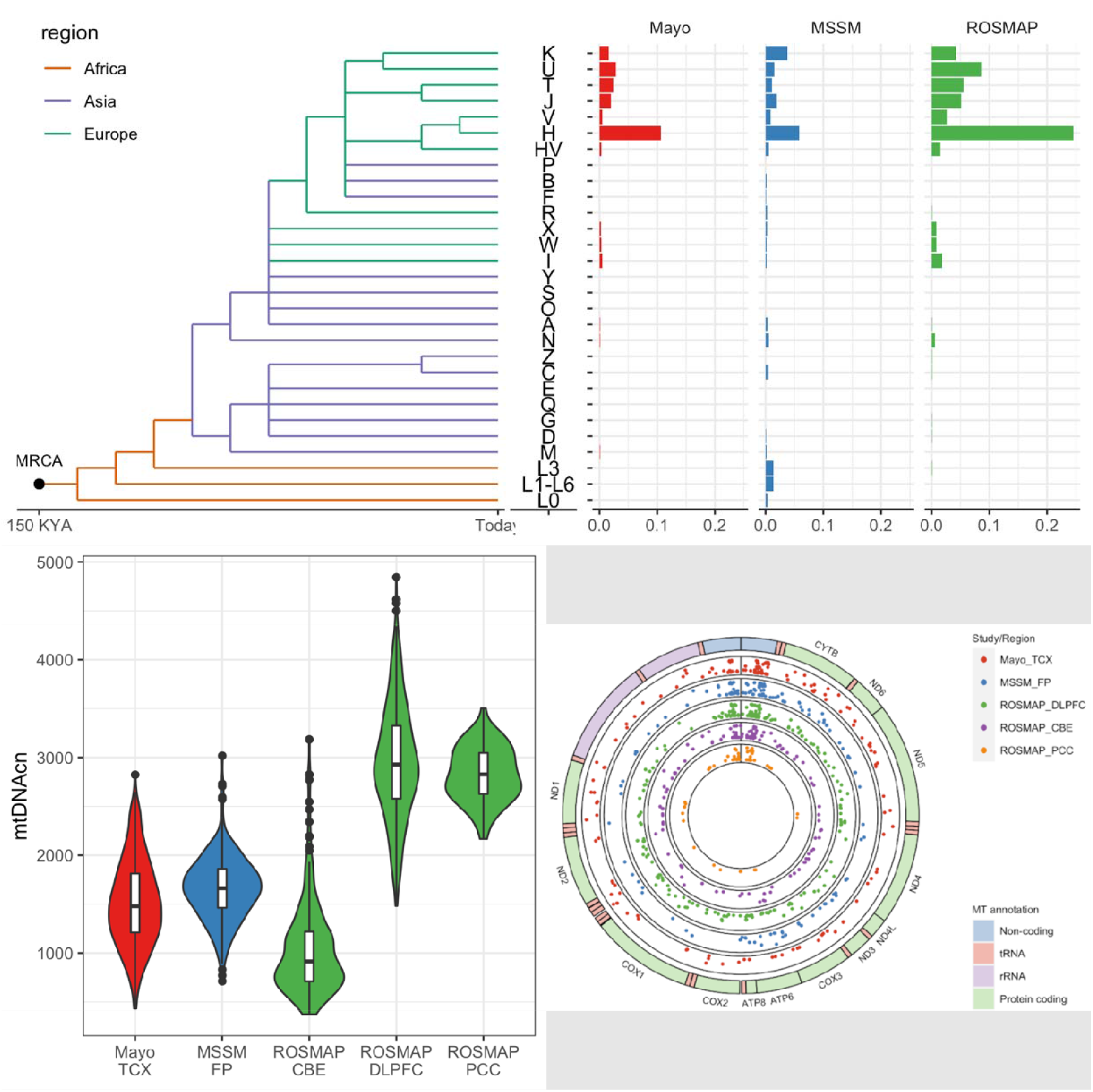
Mitochondrial genomics in AMP-AD: (Top) Simplified mitochondrial phylogenetic tree showing the evolutionary relationship between macro mitochondrial haplogroups and the proportion of haplogroups in each study. (Bottom left) Distribution of mtDNAcn across studie and source tissue. (Bottom right) Circus plot of the mitochondrial genome showing mitochondrial gene annotation on the outer circle and the average mtHz frequency at each site on the inner circles.

Given that mtDNAcn was significantly associated with AD neuropathology and cognitive performance, we used mediation analysis to evaluate if the effect of mtDNAcn levels on global cognition is mediated via AD neuropathology. Mediation determines whether the effect of mtDNAcn on global cognition decreases or disappears when AD neuropathology is included in the model. Here, we found that approximately 30% of the effect of mtDNAcn on global cognition was mediated by tangle density or global AD pathology (Table S5).

Due to the strict maternal inheritance and negligible intermolecular recombination of mtDNA, the sequential accumulation of mtDNA mutations during prehistoric human migrations has resulted in sets of specifically linked mtDNA polymorphisms in geographically defined haplotype groups or haplogroups (mtHgs) [23] (Figure 1). Genetic variation in the mitochondrial genome, either mtHg’s or individual SNPs, have been associated with AD – including mtHg J with increased risk of AD [24–26]. However, there are few definitive findings across the literature, which is characterized by contradictory findings and a lack of replication due to small sample sizes, insufficient mtDNA data, and technical challenges in data analysis [24]. In this study, we further examined the association of mtHgs with AD neuropathology and cognitive performance. We found that mtHg J was associated with increased global AD pathology, tau tangle density, and mtDNAcn. mtHg W was associated with worse semantic memory performance (Table 4, S4) and mtHg K was associated with increased neurofibrillary tangle burden (Table 3, S1).

mtDNA can carry mutations that affect all copies of the mtDNA (homoplasmic) or only a fraction of the mtDNA molecules (heteroplasmy, mtHz) within an individual cell or tissue (Figure 1). Heteroplasmy can either be transmitted maternally as low-level variants, such that both mother and offspring share the same heteroplasmic sequence deviations or occur via de novo mutations in cellular damage and aging. A higher heteroplasmy burden has been observed in brains from clinically diagnosed AD cases [27]. Additionally, increased levels of mtHz were observed in the control region of the mtDNA – the non-coding region of the mtDNA that controls replication and translation – in pathologically confirmed AD cases and were enriched in mtDNA regulatory regions [20], which has led to the hypothesis that mutations in mtDNA could influence mtDNA genome abundance. However, more recent studies found no association between mtHz and AD neuropathology [21,28]. In the present study we examined the association of the mtHz burden, across the whole mitochondrial genome and within the control region, with neuropathology burden and cognitive function (Fig 1). We found that mtHz burden across the whole genome and within the control region associated with high levels of tau tangle density (Table 4, S4, S7).

### 1.4 Study conclusions and disease implications

In this study, we evaluated the association of mtDNA abundance, mtHg’s, and mtHz burden with AD neuropathology and neurocognitive function. We found that higher mtDNA abundance robustly associated with reduced AD neuropathology burden and higher neurocognitive performance. These findings corroborate a recent report investigating the association of mtDNA quantity and quality with AD neuropathology performed in the same dataset, though using different variant calling pipelines and statistical models [28]. Validating previous findings using orthogonal methods further improves the robustness of research results across the scientific literature [29]. Our analyses also extend upon these models by evaluating the effect of mtHgs, D-loop mtHz burden, and the interactive effect of mtDNAcn and mtHz on AD neuropathology and cognitive performance. Additionally, we also examined if the effect of mtDNAcn on cognitive performance is mediated by AD neuropathology.

Given these findings, we propose an updated hypothesis for mitochondrial dysfunction in AD: that mitochondrial genome abundance – mtDNA abundance – is a relevant mechanism in AD pathogenesis. This specifies and characterizes at least one component of the proposed mitochondrially-initiated chain of events that presumably mediate a broader AD dysfunction-degeneration cascade. mtDNA abundance may also represent or reflect an important part of an individual’s genetically determined baseline mitochondrial function, influencing lifetime risk. In essence, we postulate that baseline mtDNA abundance itself contributes to baseline mitochondrial function and lifetime risk, and that propensity and sensitivity to mtDNA depletion further modulate risk, histopathology, and clinical decline. This proposal uniquely asserts the importance of mitochondrial genome abundance to AD pathogenesis. Specific mtSNPs, mtHg’s or mtHz burden may contribute, but largely through their impact on mtDNAcn or through the accentuation of mtDNAcn-related functional deficits.

Critically, the role of mtDNA abundance – and mitochondrial dysfunction more broadly – in AD also intersects with other key risk factors for Alzheimer’s disease including age, sex, ancestry, *APOE* genotype, and immunometabolism, suggesting that they may moderate the effect of mtDNA abundance on AD pathogenesis. Age is the greatest risk factor for AD, with the prevalence increasing from 5.3% in people aged 65-74 to 34.6% in people aged 85 or older [1]. Women make up nearly two-thirds of all AD cases [1]. It has been shown that mitochondrial function declines with age, and that mtDNAcn decreases after the age of 50 [30]. Although women have higher mtDNAcn levels than men, mtDNAcn increased with age in premenopausal women and decreased post-menopause [31]. In contrast, mtDNAcn levels steadily decreased with advancing age in men. As such, age associated decline in mtDNAcn would result in impaired mitochondrial function and potentially influence AD pathogenesis.

There are racial health disparities in AD with older African and Hispanic Americans two and one-and-a-half times more likely to develop AD respectively than White Americans [1]. While the higher prevalence of AD in minority populations has been attributed to disparities in medical conditions, health-related behaviors, socioeconomic risk factors - ancestry-specific genetic risk also plays a role [32]. mtDNAcn has been observed to decrease with increased discordance between mitochondrial and nuclear genetic ancestry in admixed populations. Mitonuclear discordance and corresponding lower mtDNAcn levels in admixed populations, such as African Americans and Hispanics, may contribute to the increased risk of AD observed in minority populations [33]. As such, further studies should examine the role of mitonuclear compatibility and mtDNAcn levels on AD risk in admixed populations.

*APOE*-ε4 is the strongest genetic risk factor for sporadic AD [34]. It has been associated with reduced levels of proteins involved in mitochondrial biogenesis and dynamics, oxidative stress, and synaptic integrity in post-mortem brain tissue [35]. Reduced levels of proteins involved in mitochondrial biogenesis and dynamics were further correlated with worse cognitive function [35]. *APOE*-ε4 induced human microglia-like cells (iMGLs) also show downregulation of metabolic parameters, including oxidative and glycolytic functions [36]. *APOE*-ε4 also associates with reduced mtDNAcn levels [31,37]. In summary, *APOE* also modifies mitochondrial biology and this may constitute another mechanism through which *APOE* promotes AD pathogenesis.

Recent functional genomic and pathway analyses have suggested that microglia play a central role in the etiology of AD [38]. Microglia are the innate immune cells of the central nervous system. In AD, these cells mitigate neurodegeneration through phagocytic clearance of Ab aggregates. When Ab levels accumulate beyond their clearing ability, microglia compact Ab aggregates into less toxic dense core neuritic plaques [39]. These protective activities require microglia to adopt a disease-associated microglia (DAM) state. The switch from the homeostatic to the DAM state is mediated by a mitochondrial-determined metabolic reprogramming in which the main mode of energy production changes from oxidative phosphorylation to glycolysis [40]. Mitochondria, therefore, impact microglial biology. As such, reductions in mtDNA abundance and associated mitochondrial dysfunction may impair microglial protective activities in AD pathogenesis, although this remains to be demonstrated.

### 1.5 Limitations, major challenges, and future directions

In this large study of post-mortem tissue, we find strong evidence that mtDNA abundance associates with AD neuropathology and cognitive function. However, the results of this study do not address whether mtDNA abundance causes, mediates, or is altered as a result of AD pathogenesis. A major challenge therefore remains to determine if altered mtDNAcn levels are causally related to AD pathogenesis, the specific mechanism through which reduced mtDNAcn levels influence AD pathogenesis, and if this mechanism is specific to AD or more broadly influences cellular function and the cellular response to proteinopathy. Nevertheless, there are several future research directions that may further strengthen the evidence of a causal role for mtDNA abundance and mitochondrial dysfunction in AD pathogenesis.

First, statistical genetic approaches such as polygenic risk scoring (PRS) and Mendelian randomization (MR) that leverage genomic information can be used to estimate causal associations between risk factors and disease outcomes [41]. PRS estimates an individual’s genetic propensity to a trait and can be used to infer genetic overlap between phenotypes by using the PRS of one trait to predict another. MR is a method that estimates the causal effect of an exposure on an outcome by using genetic variants as a proxy for the exposure administered as an intervention in a randomized control trial. Recent GWAS studies considered the genetic regulation of mtDNAcn in blood cells [31,37,42]. These studies could facilitate PRS and MR analyses within AD and other neurodegenerative disease datasets and assess causal relationships between mtDNAcn and AD more critically.

Second, while our overall findings suggest that mtDNA abundance in post-mortem brain tissue is associated with AD neuropathology and antemortem cognitive function, the association of mtDNAcn levels estimated from peripheral blood with AD and AD endophenotypes would provide orthogonal evidence. Peripheral blood mtDNAcn could potentially be derived from existing genotyping/sequencing data available in published cohort studies. Blood-derived mtDNAcn, however, is also affected by heterogeneous cell types driven by the relative abundance of white blood cells and platelets, which potentially limits the inferences that can be made [43]. Newly established cohorts that intend to measure blood-derived mtDNAcn should preferentially use purified cell populations or collect additional data on major cellular constituents to adjust or rule out potential confounders introduced by cell-type heterogeneity [43].

Finally, more definitive evidence of mtDNA abundance influencing AD pathogenesis would be obtained from a clinical trial of a drug that either upregulates total mitochondrial genome abundance or boosts mitochondrial mass. Peroxisome proliferator-activated receptor-gamma coactivator 1a (PGC-1a) is the master regulator of mitochondrial biogenesis and coordinates the expression of nuclear-encoded genes, including OXPHOS subunits, Tfam, and other genes involved in mtDNA gene expression [44]. Pharmacological interventions targeting PGC-1a expression have demonstrated neuroprotective effects in AD mouse models [44]. An alternative to boosting mitochondrial biogenesis is to increase the absolute levels of mtDNA independent of biogenesis. The amount of mtDNA is directly proportional to mitochondrial transcription factor A (TFAM), which facilitates the transcription and replication of mtDNA. Increased expression of TFAM in AD mouse models improved cognitive function and reduced memory impairment in aged mice [44,45]. As such, modulation of mtDNAcn through manipulation of TFAM levels may provide a therapeutic avenue to treat mitochondrial dysfunction in AD.

Similarly, ketogenic therapies represent another strategy to treat mitochondrial dysfunction in neurodegenerative diseases, including AD [46,47]. The ketogenic diet features very high-fat and low-carbohydrate intake, which triggers a shift from glucose utilization to fat-derived ketone bodies [48]. Randomized controlled trials demonstrated improved cognitive outcomes in MCI participants after ketogenic supplementation[49,50].

### 1.6 Conclusion

Mitochondrial dysfunction is one of the earliest changes observed in AD. In comparison to Aβ and tau proteinopathy, mitochondriopathy has remained of peripheral interest to much of the AD research community. Historically, the mitochondrial cascade hypothesis has not specifically emphasized the significance of mtDNA abundance on AD risk, dysfunction, and degeneration. Our study shows that reduced mtDNA abundance is associated with increased AD neuropathology and worse cognitive performance and argues that mtDNAcn-determined changes in mitochondrial function may initiate or mediate AD neuropathology or the cellular response to AD neuropathology. As such, novel therapeutic interventions that can either increase mitochondrial biogenesis or mtDNA abundance could be used to moderate AD pathogenesis.

## 2. CONSOLIDATED STUDY DESIGN AND RESULTS

Mitochondrial genomics were estimated from whole-genome sequencing data using DNA isolated from 1,249 post-mortem brain tissues obtained from three cohorts as part of the Accelerating Medicines Partnership for Alzheimer’s disease (AMP-AD) Knowledge Portal - the Religious Orders Study and the Rush Memory and Aging Project (ROSMAP), the Mount Sinai Brain Bank (MSBB), and the Mayo Clinic (Mayo) (Table 1; Figure 1; Supplementary Methods). At autopsy, neuritic plaque and neurofibrillary tangle burden were assessed using CERAD scores and Braak Stage, respectively, and donors were classified neuropathologically using the National Institute on Aging (NIA)-Reagan criteria (Supplementary Methods). In ROSMAP, additional quantitative measures of global AD pathology burden, β-amyloid load, and tau tangle density were available, along with cognitive function at each participant’s last visit prior to death across six cognitive domains, including episodic memory, perceptual orientation, perceptual speed, semantic memory, and working memory. These parameters were also used to create a measure of global cognition (Supplementary Methods).

**Table 1:** Subject Demographics

| Variable | Banner TCX, N = 60 | Mayo TCX N = 108 | MSBB FP N = 288 | ROSMAP CBE N = 265 | ROSMAP DLPFC N = 461 | ROSMAP PCC N=68 |
| --- | --- | --- | --- | --- | --- | --- |
| **Age of death**<sup>†</sup> | 81 (8) | 84 (7) | 85 (10) | 88 (7) | 89 (7) | 89 (6) |
| **APOE**<sup>‡</sup> |  |  |  |  |  |  |
| e3/e3 | 40 (67) | 58 (54) | 147 (51) | 166 (65) | 272 (59) | 40 (59) |
| e2+ | 9 (15) | 7 (6.5) | 40 (14) | 26 (10) | 67 (15) | 9 (13) |
| e4+ | 11 (18) | 43 (40) | 101 (35) | 65 (25) | 121 (26) | 19 (28) |
| **Female**<sup>‡</sup> | 25 (42) | 66 (61) | 184 (64) | 188 (71) | 301 (65) | 47 (69) |
| **PMI**<sup>†</sup> | 3 (2) | 9 (7) | 440 (327) | 8 (7) | 8 (6) | 7 (5) |
| **Braak Stage**<sup>‡</sup> |  |  |  |  |  |  |
| 0 | 0 (0) | 1 (0.9) | 13 (4.5) | 3 (1.1) | 5 (1.1) | 1 (1.5) |
| I/II | 29 (48) | 9 (8.4) | 66 (23) | 54 (20) | 65 (14) | 11 (16) |
| III/IV | 31(52) | 20 (19) | 82 (28) | 144 (54) | 245 (53) | 39 (57) |
| V/VI | 0(0) | 77 (72) | 127 (44) | 64 (24) | 145 (32) | 17 (25) |
| **CERAD Score**<sup>‡</sup> |  |  |  |  |  |  |
| None | - | - | 109 (38) | 80 (30) | 159 (35) | 24 (35) |
| Sparse | - | - | 93 (32) | 82 (31) | 170 (37) | 23 (34) |
| Moderate | - | - | 27 (9.4) | 18 (6.8) | 49 (11) | 3 (4.4) |
| Frequent | - | - | 27 (9.4) | 18 (6.8) | 49 (11) | 3 (4.4) |
| **ADNC**<sup>‡</sup> |  |  |  |  |  |  |
| Control | 60(100%) | 31(29%) | 105(36%) | 114(43%) | 147(32%) | 23(34%) |
| Case | 0(0%) | 77(71%) | 183(64%) | 151(57%) | 313(68%) | 45(66%) |

<sup>†</sup> n (%); <sup>‡</sup> mean (sd)
TCX: Temporal Cortex; FP: Frontal pole; CBE: Cerebellum; DLPFC: Dorsolateral prefrontal cortex; PCC: Posterior cingulate cortex.

To investigate the association of mtDNAcn, mtHg, and mtHz on ADNC, Braak staging, and CERAD scores, we conducted a joint analysis of the three AMP-AD cohorts adjusting for *APOE*, age of death, post-mortem interval, sex, study, and tissue. Increased mtDNAcn levels were associated with reduced odds of neuropathological AD, (OR [95% CI]: 0.64 [0.48, 0.84] p=0.002) and reduced neurofibrillary tangle burden (OR [95% CI]: 0.61 [0.48, 0.78], p= 7.78E-05), but not with neuritic plaque burden (Table 3, S1). Additionally, we found that mtHz burden was not associated with AD neuropathology (Table 3, S1), and there was no significant interaction between mtDNAcn and mtHz burden (Table S2).

**Table 2:** Mitochondrial genomics across sample/tissue

| Variable | Banner<br>TCX,<br>N = 60 | Mayo TCX<br>N = 108 | MSBB FP<br>N = 288 | ROSMAP<br>CBE<br>N = 265 | ROSMAP<br>DLPFC<br>N = 461 | ROSMAP PCC<br>N=68 |
| --- | --- | --- | --- | --- | --- | --- |
| **mtDNAcn**<sup>‡</sup> | 1424(405) | 1416(349) | 1653(319) | 1041(469) | 2956(592) | 2830(279) |
| **mtHz**<sup>†</sup> |  |  |  |  |  |  |
| 0 | 6 (11) | 7 (6.8) | 16 (8.2) | 96 (40) | 26 (6.2) | 4 (6.2) |
| 1 | 20 (37) | 38 (37) | 76 (39) | 78 (33) | 124 (30) | 24 (38) |
| 2 | 15 (28) | 32 (31) | 60 (31) | 47 (20) | 153 (37) | 21 (33) |
| 3 | 7 (13) | 12 (12) | 27 (14) | 13 (5.4) | 72 (17) | 10 (16) |
| 4 | 3 (5.6) | 12 (12) | 12 (6.2) | 3 (1.3) | 33 (7.9) | 2 (3.1) |
| 5 | 2 (3.7) | 1 (1.0) | 2 (1.0) | 0 (0) | 8 (1.9) | 1 (1.6) |
| 6 | 1 (1.9) | 1 (1.0) | 1 (0.5) | 2 (0.8) | 1 (0.2) | 2 (3.1) |
| 9 | 0 (0) | 0 (0) | 0 (0) | 0 (0) | 1 (0.2) | 0 (0) |
| **mtHg**<sup>†</sup> |  |  |  |  |  |  |
| H | 26 (48) | 54 (52) | 71 (37) | 111 (46) | 191 (46) | 21 (33) |
| I | 1 (1.9) | 3 (2.9) | 3 (1.5) | 5 (2.1) | 18 (4.3) | 2 (3.1) |
| J | 5 (9.3) | 11 (11) | 22 (11) | 17 (7.1) | 40 (9.6) | 7 (11) |
| K | 5 (9.3) | 8 (7.8) | 51 (26) | 14 (5.9) | 32 (7.7) | 8 (12) |
| T | 9 (17) | 9 (8.7) | 10 (5.2) | 31 (13) | 34 (8.1) | 9 (14) |
| U | 4 (7.4) | 12 (12) | 19 (9.8) | 42 (18) | 71 (17) | 9 (14) |
| V | 1 (1.9) | 3 (2.9) | 11 (5.7) | 10 (4.2) | 22 (5.3) | 5 (7.8) |
| W | 1 (1.9) | 2 (1.9) | 3 (1.5) | 2 (0.8) | 5 (1.2) | 3 (4.7) |
| X | 2 (3.7) | 1 (1.0) | 4 (2.1) | 7 (2.9) | 5 (1.2) | 0 (0) |
| **Neurons**<sup>‡</sup> | 0.069<br>(0.016) | 0.063<br>(0.015) | 0.060<br>(0.026) | 0.069<br>(0.022) | 0.061<br>(0.024) | 0.069 (0.019) |
| **Astrocytes**<sup>‡</sup> | 0.008<br>(0.006) | 0.007<br>(0.005) | 0.036<br>(0.019) | 0.044<br>(0.012) | 0.049<br>(0.013) | 0.045 (0.011) |

<sup>†</sup> n (%); <sup>‡</sup> mean (sd)
TCX: Temporal Cortex; FP: Frontal pole; CBE: Cerebellum; DLPFC: Dorsolateral prefrontal cortex; PCC: Posterior cingulate cortex.

**Table 3:**
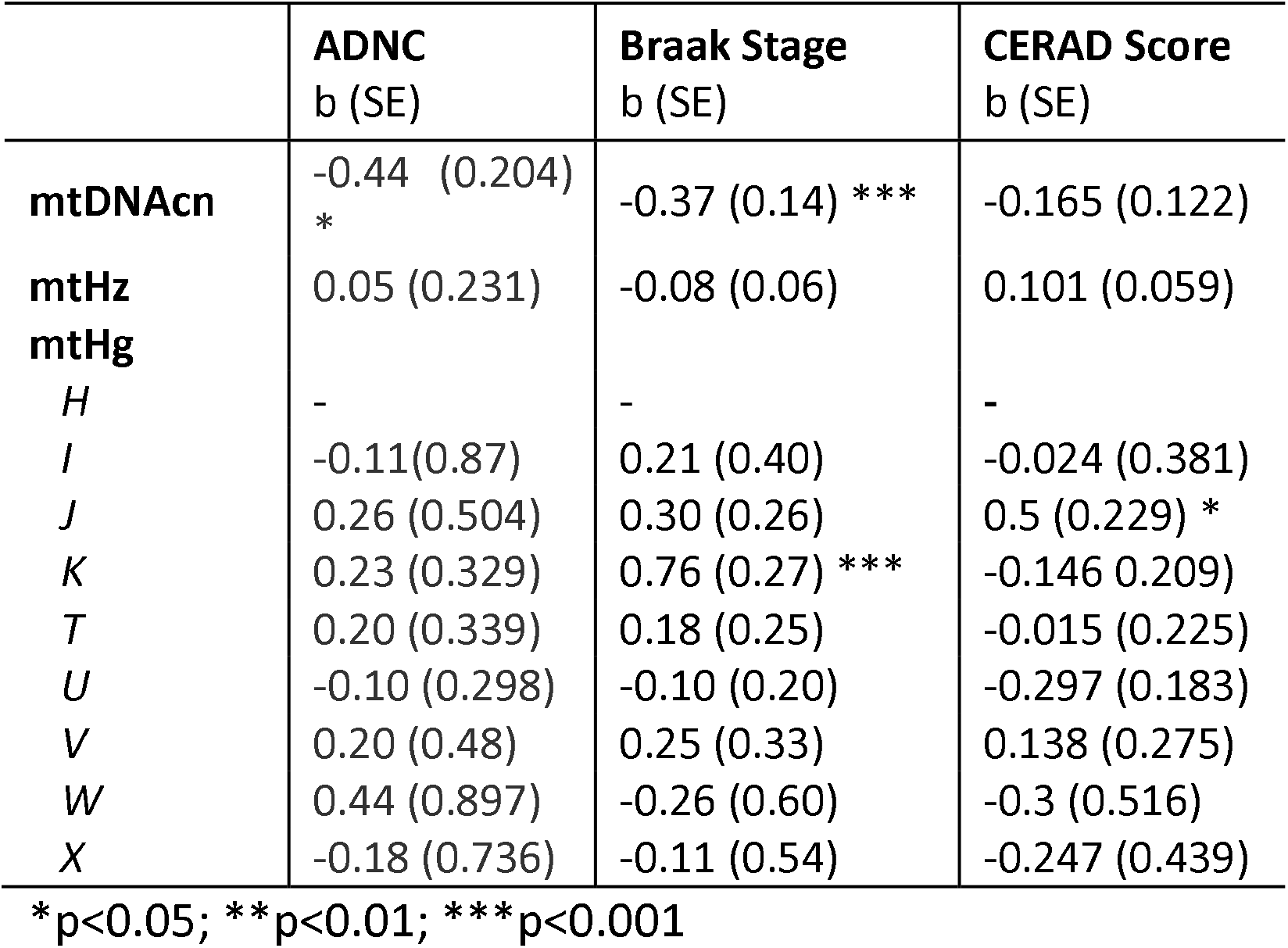
Association of mtDNAcn, mtHg’s, and mtHz burden with Alzheimer’s disease neuropathology in AMP-AD

To further understand the relation of mtDNA heteroplasmy with AD neuropathology, we examined the association of mtHz burden limited to the control region as mtDNA replication is initiated in the control region (Fig. 1), and heteroplasmy burden in this region has been previously associated with AD [25]. We observed no significant associations (Table S3). Next, we tested the association of mtHg with AD neuropathology and found that mtHg J was associated with increased amyloid burden (OR [95% CI]: 1.65 [1.05, 2.58] p= 0.028) and with increased mtDNAcn (Table S11) while mtHg K was associated with high levels of neurofibrillary tangle burden (Table 3, S1). No other mtHg was associated with AD neuropathology.

Among 686 participants from the ROSMAP cohort, we performed multivariable linear regression to investigate the association between mtDNAcn, mtHg, and mtHz on post-mortem amyloid-β and tau burden and antemortem cognitionGi (Table 4, S4). Given that mtDNAcn was significantly associated with AD neuropathology and cognitive performance, we further used mediation analysis to evaluate if the effect of mtDNAcn levels on global cognition is mediated via AD neuropathology. The effect of mtDNAcn on global cognition was partially mediated by tangle density and global AD pathology, accounting for 27% and 28% of the total effects, respectively (Table S5). These findings suggest that mtDNAcn levels influence cognitive impairment partially through AD neuropathology.

**Table 4:**
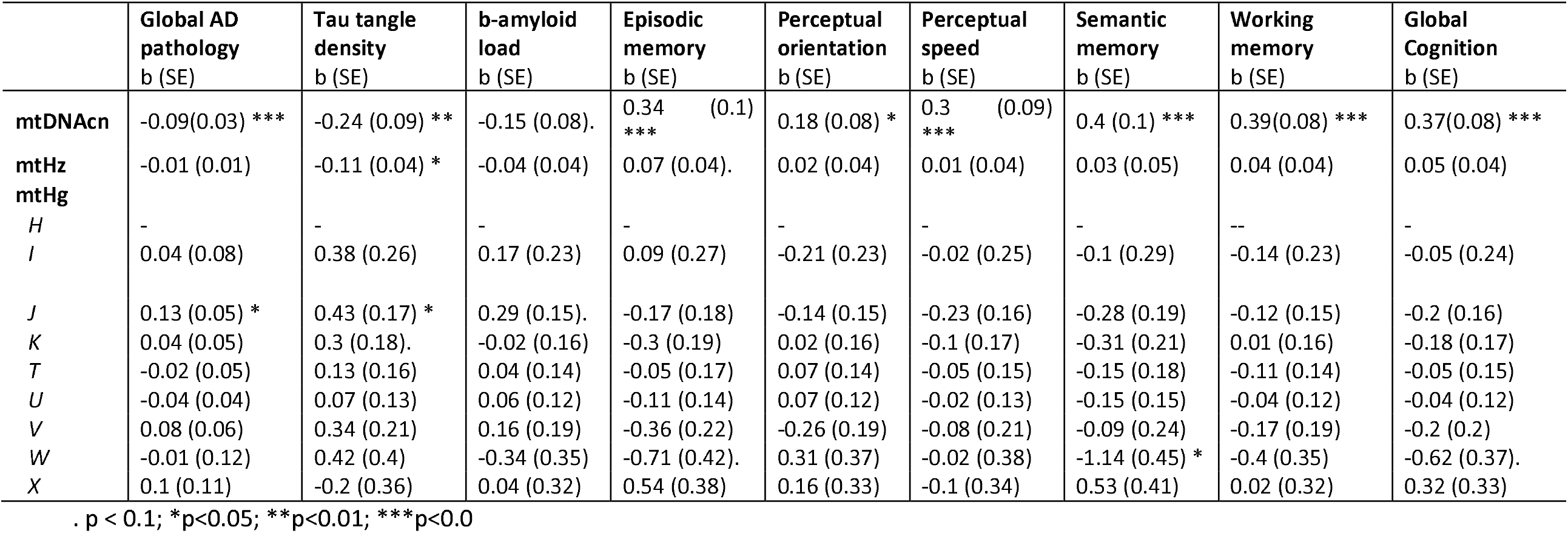
Association of mtDNAcn, mtHg, mtHz with AD neuropathology and cognitive function in ROSMAP

Next, we used the ROSMAP dataset to examine the association between mtHz across the whole mitochondrial genome and the D-loop on post-mortem amyloid-β and tau burden and antemortem cognition. We found that mtHz burden across the whole mitochondrial genome and restricted to the D-loop was associated with tau tangle density (Table 4, S4, S7). The interaction between total mtHz and mtDNAcn was not significantly associated with cognitive performance or AD neuropathology (Table S6). Likewise, we tested whether haplogroups were associated with amyloid-β and tau burden and antemortem cognition. Indeed, we found that mtHg J associated with increased global AD pathology, tau tangle density, and mtDNAcn, while mtHg W associated with worse semantic memory performance (Table 4, S4).

Neurons have a high number of mitochondria, and the proportion of neurons changes over the course of AD. For these reasons mtDNAcn estimates from bulk brain tissue can be confounded by cell type heterogeneity. To account for cell-types, we estimated neuron and astrocyte proportions in a subset of participants using bulk tissue RNA-seq data obtained from the same brain regions from which the DNA for WGS was isolated. Next, we re-ran the regression models adjusting for *APOE*, age of death, post-mortem interval, sex, study, tissue, and neuron-astrocyte enrichment. In AMP-AD (n = 855), higher mtDNAcn remained associated with reduced tau burden (b (SE) = 0.69 (0.15), p= 0.017) (Table S8). In ROSMAP (n = 197), higher mtDNAcn remained significantly associated with reduced global AD pathology and better cognitive performance in domains associated with working memory and perceptual speed (Table S9. Loss of significance across other outcomes may reflect inadequate statistical power due to smaller sample sizes (Table S10).

The effect of mitochondrial dysfunction on the risk of AD could be modulated by other factors, such as *APOE*-ε4 locus, age, and sex. We therefore sought to examine the associations of these risk factors with mtDNAcn and mtHz in the AMP-AD datasets. *APOE*-ε4 was associated with lower levels of mtDNAcn (β (SE) = - 0.09 (0.04), p= 0.017) (Table S11). We found no significant association between mtDNAcn and advanced age. mtHg J was associated with increased mtDNAcn levels (β (SE) = 0.15 (0.05), p= 0.01) (Table S11). In contrast to the mtDNAcn, heteroplasmy levels were not associated with *APOE*-ε4 but were associated with age (β (SE) = 0.03 (5.46), p= 5.93E-08) (Table S11). In addition, we found a significant association between heteroplasmy levels with several mtHgs, including mtHg I, mtHg K, mtHg T, mtHg U, mtHg V, and mtHg W (Table S11). Sex did not associate with mtDNAcn or heteroplasmy burden.

## 3. DETAILED METHODS

### 3.1 Religious Orders Study and the Memory and Aging Project

#### Neuropathological characterization

The Religious Orders Study (ROS) and the Memory Aging Project (MAP) are two prospective longitudinal cohort studies of aging and dementia. Participants of ROS are enrolled as older Catholic nuns, brothers, and priests whilst MAP enrolls older community-dwelling adults. Both studies include persons without known dementia who consent for annual clinical assessments and brain, spinal cord, and muscle donation after death. Collectively, ROSMAP contains harmonized clinical and post-mortem data from both ROS and MAP, including annual clinical and cognitive assessments, neuropathological traits, and biomarkers assessed using brain autopsies, whole-genome sequencing, and richly phenotyped multi-omics [51,52].

#### Neuropathology Endophenotypes

Neuropathological endophenotypes from ROSMAP we examined include beta-amyloid burden, diffuse plaque burden, neuritic plaque burden, neurofibrillary tangle burden, tau tangle density, global AD pathology burden, CERAD Score, Braak Stage, and NIA-Reagan neuropathological diagnosis [52]. Diffuse plaque burden, neuritic plaque burden, and neurofibrillary tangle burden were determined by microscopic examination of modified Bielschowsky silver-stained slides from five regions, namely midfrontal cortex, midtemporal cortex, inferior parietal cortex, entorhinal cortex, and hippocampus. Composite scores for these neuropathological markers were computed by averaging the scores obtained from five brain regions: midfrontal cortex, midtemporal cortex, inferior parietal cortex, entorhinal cortex, and hippocampus [52]. Subsequently, semi-quantitative measurements of neuritic plaque density and Neurofibrillary tangle burden were determined. As recommended by the Consortium to Establish a Registry for Alzheimer’s Disease (CERAD), neuritic plaque density is modified to be implemented without adjustment for age and clinical diagnosis [53]. Neurofibrillary tangle burden was measured using Braak Staging, a semiquantitative measure of severity of neurofibrillary tangle (NFT) pathology [54]. A neuropathological diagnosis of AD was based on modified NIA-Reagan criteria that emphasize CERAD and Braak staging. The neuropathological evaluation is conducted blinded to clinical information, including a diagnosis of dementia. Participants who had an intermediate to high likelihood of AD according to the NIA-Reagan criteria were classified as having a pathological diagnosis of AD. Global AD pathology burden was calculated as a quantitative summary of AD pathology derived from three AD pathologies: neuritic plaques, diffuse plaques, and neurofibrillary tangles (quantified as described above). Counts of each pathology were standardized and averaged across brain regions to obtain a single score for the global AD pathology burden. Amyloid plaque and tau tangle pathology were identified by molecular-specific immunohistochemistry using anti-human antibodies and quantified by image analysis. The percentage of cortex occupied by amyloid-beta plaques and tau tangles in eight regions was recorded, namely hippocampus, entorhinal cortex, midfrontal cortex, inferior temporal cortex, angular gyrus, calcarine cortex, anterior cingulate cortex, and superior frontal cortex. Composite scores for overall amyloid-β burden and tau tangle density were computed by averaging scores from the eight brain regions. Global AD pathology, amyloid burden, and tau tangle density were square rooted for better statistical properties.

#### Cognitive Endophenotypes

ROSMAP participants undergo a series of neuropsychiatric tests at each visit, with related tests used to construct composite measures across five cognitive domains including episodic memory, perceptual orientation, perceptual speed, semantic memory, working memory, and global cognition [52]. Each participant’s last cognitive test prior to death was used in downstream analyses.

#### Whole-genome sequencing

DNA for WGS was extracted from the dorsolateral prefrontal cortex (DLPC, n = 466), posterior cingulate cortex (PCC, n = 68), and cerebellum (CB, n = 265) [51] Before quality control, we preferentially kept samples with DNA isolated from dorsolateral prefrontal cortex (DLPFC) > posterior cingulate cortex (PCC) > cerebellum (CER) > whole blood > peripheral blood mononuclear cells (PBMCs) > other (n = 805).

#### Exclusion Criteria

Samples were excluded based on not passing WGS quality control (n =16), non-European ancestry, (n=1), belonging to a non-European haplogroup (n= 62), and phenotype completeness. A total of 785 brain samples from DLPFC, PCC, and CER were retained for downstream analysis.

### 3.2 Mayo clinic

#### Neuropathological characterization

The Mayo Clinical case-control cohort is composed of post-mortem brain tissue obtained from the Mayo Brain Bank (n = 108) or the Banner Sun Health Institute (n = 60) [55]. Samples were classified as either control, pathologic aging, AD, or progressive supranuclear palsy. Controls had a clinical dementia rating score of 0 and a Braak Stage of < 3; pathological aging had a Braak stage of <=3, CERAD score of >=2, and lacked any other pathological diagnoses. AD cases were diagnosed as possible or probable AD according to NINCDS-ADRDA criteria and had a Braak Stage of > 4. Since age at death was right-censored at 95 years for HIPPA compliance, age of death was treated as a winsorized variable. DNA for WGS was extracted from the temporal cortex (n = 341).

#### Exclusion Criteria

We used the same quality control procedures as the ROS/MAP WGS data to exclude samples that failed quality control (n = 0), were missing phenotypes (n =0) did not have European ancestry (n = 0) and were not European haplogroups (n = 24). Additionally, we excluded samples diagnosed with progressive supranuclear palsy (n =86). After quality control, 168 samples with DNA isolated from the cerebellum were retained for downstream analysis.

### 3.3 Mount Sinai Brain Bank

#### Neuropathological characterization

The Mount Sinai Brain Bank (MSBB) is a case-control cohort composed of post-mortem brain tissue from 364 brains and has been previously described [56]. This cohort was assembled after applying stringent inclusion/exclusion criteria that represents the full spectrum of cognitive and neuropathological disease severity in the absence of discernable non-AD pathology. Amyloid plaque burden was determined through semiquantitative estimates of neuritic plaque density as recommended by CERAD, as modified to be implemented without adjustment for age and clinical diagnosis [53]. NFT burden was determined through semiquantitative Braak Staging [54]. A neuropathological diagnosis of AD was based on modified NIA-Reagan criteria that emphasize CERAD and Braak staging, with the neuropathological evaluation conducted blinded to clinical information and a diagnosis of dementia. Participants who had an intermediate to high likelihood of AD according to the NIA-Reagan criteria were classified as having a pathological diagnosis of AD. DNA for whole-genome sequencing was isolated from post-mortem brain tissue dissected from the frontal pole (Brodmann area 10).

#### Exclusion Criteria

We followed the same approach to quality control as was used for the previous WGS data, including filtering on the basis of data completeness, WGS quality control, European ancestry, and European haplogroups. After quality control, 288 samples were retained for downstream analyses.

### 3.4 Mitochondrial variant calling

#### Whole-genome sequencing and nuclear variant calling

Whole-genome sequencing library preparation and variant calling for ROSMAP [52], MSBB [56] and Mayo [55] are available through the AMP-AD Knowledge Portal (syn26243166). Briefly, whole-genome sequencing (WGS) libraries were prepared using the KAPA Hyper Library Preparation Kit in accordance with the manufacturer’s instructions. Libraries were sequenced on an Illumina HiSeq X sequencer (v2.5 chemistry) using 2 × 150bp cycles. Whole Genome data are processed on an NYGC automated pipeline, aligned to the GRCh37 human reference using the Burrows-Wheeler Aligner (BWA-MEM v0.7.08), and processed using the GATK best-practices workflow.

#### Mitochondrial DNA Variant calling

Mitochondrial homoplasmic and heteroplasmic variants were called following GATK’s Best Practice Mitochondrial Pipeline [57] implemented as a Snakemake workflow [58] (https://github.com/marcoralab/gatk-mitochondria-pipeline). GATK v4.2.0 was used to run the pipeline with the rCRS as the mitochondrial reference sequence. Sequence reads mapped to the mtDNA were extracted from .bam files, converted into FASTQ files, aligned to the rCRS and to the rCRS shifted by 8000 base pairs using BWA-MEM v0.7.17, and converted back into .bam files. Mitochondrial variants were subsequently detected using the mitochondrial mode of GATK’s MuTect2 variant caller, which is designed to account for higher coverage and potential low heteroplasmy variants. Variants called from the standard rCRS and shifted rCRS were merged into one VCF file. The shifted reference is used for detecting variants around the artificial start/end position of the mitochondrial genome (coordinates chrM:16025-16569 and chrM:1-575), and the standard reference is used for identifying variants on the rest of the mitochondrial genome (bp 576-16024). Variants showing weak evidence or strand bias were then filtered, with the median autosomal chromosome coverage estimated using Mosdepth (v0.3.2) [59] to filter potential polymorphic NUMT variants. We also accounted for possible mtDNA contamination by using Haplochecker (v1.3.2) [60], a minimum variant allele frequency of 0.01, and an F-score beta of 1. Indels were left-aligned, and multiallelic sites were split. We denote variants with a variant allele frequency (VAF) of 0.03-0.95 as heteroplasmic and variants with a VAF of 0.95-1.00 as homoplasmic. Individual VCF files were combined, variant annotations applied, and variant and sample filtered using Hail [57].

#### Haplogroup assignment

Haplocheck was used to assign mitochondrial haplogroups [60] based on Phylotree 17.

#### Mitochondrial Heteroplasmy Burden

mtHz burden was defined as the number of heteroplasmic variants per individual.

#### Estimation of mitochondrial genome abundance from WGS data

The average sequence coverage of the autosomal chromosomes (cov_n_) and of the mitochondrial genome (cov_mt_) was calculated using Mosdepth from .bam files [59]. mtDNAcn was defined as (cov_n_/cov_mt_) × 2.

### 3.5 Estimation of neuronal cell proportion from RNA-seq data

Bulk tissue RNAseq data were processed, and quality controlled as previously described by the RNAseq Harmonization Study (syn9702085), and the transcript abundance was downloaded from the AMP-AD knowledge portal (ROSMAP: syn10507749, MSBB: syn10507727; Mayo: syn10507725). Gene level transcripts per million (TPM) were generated from the transcript abundance and were used by xCell to estimate cell type proportions [61,62], only allowing for deconvolution of astrocytes and neurons.

### 3.6 Statistical analysis

All statistical analysis was performed with R and implemented using Snakemake workflows.

#### Association of mtDNAcn, mtHg’s and mtHz burden with ADNC, Braak Staging, and CERAD Staging

A joint analysis of the three AMP-AD cohorts was conducted to investigate the association of mtDNAcn, mtHg, and mtHz on ADNC, Braak staging, and CERAD scores. Multivariable logistic regression analysis was used to investigate the association of mtDNAcn, mtHg, and mtHz with ADNC (n = 1249; ROSMAP (n=793), MSBB (n=288), Mayo (n=167)) adjusting for *APOE*, age of death, post-mortem interval, sex, study, and tissue. The association of mtDNAcn, mtHg, and mtHz with Braak staging (n = 1249; ROSMAP (n=793), MSBB (n=288), Mayo (n=168)) and CERAD scores (n = 1081; ROSMAP (n=793), MSBB(n=288)) was examined using ordinal logistic regression adjusting for *APOE*, age of death, post-mortem interval, sex, study, and tissue. As absolute mtDNAcn levels moderate the effect of heteroplasmy burden, we also investigated the interaction between mtDNAcn and mtHz by introducing an interaction term in the above models.

#### Association of mtDNAcn, mtHg, and mtHz burden with post-mortem amyloid plaques, tau tangles, and ante mortem cognition in ROSMAP

In ROSMAP (n = 686) we further investigated the association of mtHg’s, mtHz, and mtDNAcn with tau postmortem tangle density, amyloid plaques, and ante mortem episodic memory, perceptual orientation, perceptual speed, semantic memory, working memory, and global cognitive performance. Multivariable linear regression was used to investigate the association of mtDNAcn, mtHg, and mtHz with amyloid plaques, tangles, and cognition adjusting for age of death, *APOE*, post-mortem interval, sex, tissue, and education for cognitive outcomes. We further evaluated the interaction between mtDNAcn and mtHz by introducing an interaction term into the models. Additionally, we counted the heteroplasmy in the mtDNA control region (coordinates chrM:16025-16569 and chrM:1-575) and evaluated associations with post-mortem amyloid plaques, tangle burden, and ante mortem cognition.

#### Sensitivity analyses adjusting for neuronal cell proportion

mtDNAcn levels estimated from bulk tissue can be influenced by cell type proportion. As neurons have a relatively high number of mitochondria and the proportion of neurons changes during the course of AD, mtDNAcn levels can be confounded by neuronal loss. We therefore further evaluated the association of the mtDNAcn with ADNC, Braak staging, and CERAD scores in AMP-AD adjusting for neuronal and astrocyte cell proportions estimated from bulk tissue RNAseq data, RNA integrity number (RIN), and batch. Regression models were conducted separately in each cohort and a fixed effects meta-analysis was used to obtain an overall effect of mtDNAcn on AD neuropathology. In ROSMAP, we also evaluated the association between mtDNAcn and amyloid, tau, and cognition adjusting for neuronal and astrocyte cell proportions, RIN, and batch. The sample sizes for these analyses were smaller due to requiring samples to have whole genome sequencing and RNA sequencing from the same brain region (n = 855; ROSMAP (n =794), MSBB (n=172), Mayo (n=107)).

#### Mediation analysis of mtDNAcn effect on global cognition by AD neuropathology

Mediation analysis was performed by first estimating the effect of mtDNAcn on global cognition at last assessment using linear regression adjusting for mtHz burden, mtHgs, age of death, *APOE*, post-mortem interval, sex, tissue, and AD neuropathology – amyloid plaques, tangle density, or global pathology. The mediator model was constructed looking at the association of mtDNAcn with each AD neuropathology and adjusting for the same covariates. Mediation analysis was then used to estimate the proportion of risk in the outcome model explained by a direct effect of mtDNAcn on global cognition —the average direct effect (ADE) — and the proportion that was mediated by AD neuropathology — the average causal mediation effect (ACME). Confidence intervals were estimated using resampling (10,000 empirical bootstraps). Mediation analyses were performed with the mediation package [63] in R.

#### Association of APOE genotype with mtDNAcn levels

We evaluated the association of *APOE* genotype with mtDNAcn levels using linear regression adjusting for age, sex, PMI, haplogroups, disease status, and brain tissue stratified by study in the AMP-AD datasets.

## Data Availability

All data produced in the present study are available upon reasonable request to the authors

## Supplementary Tables

Table S1: Full model of association of mtDNAcn, mtHg’s, and mtHz burden with Alzheimer’s disease Neuropathological change in AMP-AD

Table S2: Interactive effect of mtDNAcn and mtHz burden on Alzheimer’s disease Neuropathological change in AMP-AD

Table S3: Association of heteroplasmy burden in the mtDNA control region with Alzheimer’s disease neuropathology in AMP-AD

Table S4: Full model of Association of mtDNAcn, mtHg, mtHz with AD neuropathology and cognitive function in ROSMAP

Table S5: Mediation analysis

Table S6: Interactive effect of mtDNAcn and mtHz burden on AD neuropathology and cognitive function in ROSMAP

Table S7: Association of heteroplasmy burden in the mtDNA control region with AD neuropathology and cognitive function in ROSMAP

Table S8: RNA-seq sensitivity analysis AMP-AD

Table S9: Adjusted RNA-seq sensitivity analysis ROSMAP

Table S10: Unadjusted RNA-seq sensitivity analysis ROSMAP

Table S11: Association of age and *APOE* with mtDNAcn levels and mtHz burden

